# A Transdiagnostic Dimensional Approach to Behavioral Dysregulation: Examining Reward and Punishment Sensitivity Across Psychopathology

**DOI:** 10.1101/2024.10.14.24315505

**Authors:** Taiki Oka, Akihiro Sasaki, Nao Kobayashi

## Abstract

**Aim:** Theoretically, deficits in reward/punishment sensitivity are considered an essential component associated with behavioral dysregulation, which is characteristic of compulsive-impulsive disorders. However, recent studies have indicated that several disorders are linked to problems with reward/punishment sensitivity, and their results have been inconsistent. This lack of clinical specificity and robustness might reflect more general problems with traditionally diagnostic categories of psychiatry. To address these concerns, we investigated whether a transdiagnostic dimensional approach could more effectively examine clinical associations related to reward/punishment sensitivity for behavioral dysregulation.

**Methods:** Using multiple psychiatric symptom scores and reward/punishment sensitivity in online general-population samples (N = 19505), we applied factor analyses to extract transdiagnostic symptom dimensions. Then, we conducted a mixed-effect generalized linear model to examine the relationships between psychopathology and reward/punishment sensitivity.

**Results:** We extracted three transdiagnostic dimensions, which were validated between two separate datasets: ‘Compulsive hypersensitivity (CH), ‘Social withdrawal (SW),’ and ‘Addictive behavior (AB).’ While SW was associated with reward sensitivity negatively and punishment sensitivity positively, AB showed opposite associations. On the other hand, CH was positively associated with both sensitivities.

**Conclusion:** Our results highlight the importance of reward/punishment sensitivity for our understanding of behavioral dysregulation, especially in the compulsive-impulsive dimension. Moreover, these findings underscore how transdiagnostic perspectives contribute to a more powerful examination of reward/punishment deficits than studies focusing on a categorical disorder.

## Introduction

A loss of control over repetitive self-destructive behaviors is problematic and found in a variety of disorders, particularly obsessive-compulsive and addictive disorders^1,2^. Moreover, with the advent of digital technological devices, a new psychiatric problem has emerged, such as gaming disorder, which characterizes the maladaptive engagement in game playing^3^. Researchers have suggested that issues with processing rewards and/or punishments may be involved in the characteristic of such behavioral dysregulation^4^, e.g., insensitivity to punishment in compulsive tobacco abuse and high sensitivity to reward in obsessive-compulsive disorder (OCD)^5,6^. In addition, problems related to reward/punishment sensitivity have been shown to be associated with abnormalities in brain functions such as the amygdala^7^, suggesting that understanding the relationship may be a promising target for developing new drug and psychotherapeutic interventions for behavioral dysregulation.

Importantly, the specificity of the association between reward/punishment sensitivity and each diagnosis of psychopathology has not been established. Indeed, similar deficits in reward/punishment sensitivity have been reported in a number of other patient groups, including internalizing disorders (depression, anxiety, etc.)^8^. Given that such disorders are not characterized by repetitive, compulsive behavior and impulse control deficits, it is questionable to hypothesize that reward/punishment sensitivity is a neuropsychological mechanism involved only in behavioral dysregulation associated with specific obsessive/addictive diagnoses. Moreover, the results are not consistent across studies. For example, with regard to addiction, some papers reported that symptoms were positively associated with reward sensitivity^9,10^, while others suggested more complex relationships with reward/punishment sensitivity^11–13^. Unfortunately, this lack of specificity and consistency is ubiquitous in psychiatric research^14^. This problem may be the result of a broader problem in which conventional psychiatric diagnostic labeling does not reflect a neurobiologically or psychologically beneficial phenomenon. Of particular relevance to the present study is the suggestion that behavioral preoccupations such as obsessive-compulsive and gaming disorders need to be viewed broadly as disorders of behavioral control related to compulsive/impulsive spectrum^4^, and more broadly, that the majority of patients diagnosed with these disorders meet lifelong depression and anxiety or meet other criteria^15,16^. Moreover, compulsive-impulsive spectrum has recently been suggested as a theoretical transdiagnostic aspect representing a loss of control over repetitive self-destructive behaviors across various disorders^1,2^. Given these perspectives, current studies following categorical-based diagnoses may struggle to disentangle the specific neurocognitive and psychological mechanisms of behavioral dysregulation for each diagnostic category.

One such proposed approach beyond diagnosis is the Research Domain Criteria (RDoC), which helps identify transdiagnostic and translational features of affective disorders^17^. Mechanistic clarification using this framework has been accomplished to some extent by studies examining separable patient clusters within groups diagnosed with the same disorder^18,19^. However, identifying robust, generalizable, specific markers that contribute to psychiatric comorbidity is limited by the small sample sizes typical of patient studies^20^. Previous studies have, therefore, used a transdiagnostic dimensional approach that leverages the efficiency of large-scale online data collection from healthy subjects rather than diagnosed patients to identify associations between transdiagnostic dimensions and various behavioral properties^21,22^. The use of such data-driven approaches to uncover transdiagnostic psychological mechanisms has become a major trend in recent psychiatric research^23,24^. However, few studies have focused on and conducted on the transdiagnostic levels of the obsessive-compulsive/impulsive spectrum and reward/punishment sensitivity.

The present study addressed this issue using an unsupervised approach based on a large online sample of approximately 20,000 individuals. Initially, we attempted to extract transdiagnostic factors based on data from eight questionnaires related to behavioral dysregulation and psychopathology. We then analyzed the data using generalized linear regression models to identify specific/heterogeneous associations between each disorder/dimension and reward/punishment sensitivity. As a result, three factors characterized by compulsive hypersensitivity, social withdrawal, and addictive disorder were extracted, and each was found to have a transdiagnostic association with reward/punishment sensitivity. The results of this study are a step forward toward establishing a transdiagnostic approach such as RDoC and suggest that dimensional markers of psychiatric disorders may correspond closely to neuropsychological characteristics.

## Methods

### Ethics

This study was approved by the Ethics Committee of the Advanced Telecommunications Research Institute International (Japan) (No.182H). All participants gave informed consent before responding to the surveys.

### Participants and procedure

All procedures were approved by the Ethics Committee of the Advanced Telecommunications Research Institute International (Japan) (approval No.182H). In September 2023, we collected questionnaire data from 20,000 respondents monitored by Macromill Survey Services. Of these, 495 participants were excluded due to the identification of unreliable responses, e.g., they responded identically to all items using only the maximum or minimum values in the questionnaires, which included reversed questions. As a result, 19,505 participants were included in the analysis.

### Measures

#### Reward/punishment sensitivity questionnaire

We assessed the reward/punishment sensitivity score using a behavioral inhibition/approach system questionnaire (BIS/BAS^25^). The BIS/BAS has been defined as a subscale of the affective valence system characterized as reward (positive) punishment (negative) sensitivity in RDoC, which may better capture aspects of reward/punishment sensitivity across psychiatric disorders. Although the BAS is generally considered to be divided into three sub-factors (BAS-Drive, BAS Reward Responsiveness, and BAS FunSeeking), the previous article indicated that it has a one-factor structure, so for this analysis, the entire BAS was considered as a variable^26^.

#### Self-report psychiatric questionnaires

Participants completed self-report questionnaires assessing attentional/hyperactivity disorder (ADHD) based on ASRS^27^; social anxious symptoms based on LSAS^28^; impulsivity based on BIS^29^; OCD obsessive-compulsive disorder based on OCI^30^; psychological distress general mental health based on K6^31^; alcohol-related disorder and tobacco dependence based on CAGE^32^/TDS^33^; gaming disorder based on POGQ^34^. These domains are considered to be closely interrelated given the comorbidity and the interrelationship^35,36^. In the following regression analyses, they were analyzed based on each questionnaire unit.

### Analysis

#### Factor analysis

To obtain a parsimonious latent structure for explaining shared variance at the item level across questionnaires, factor analysis with maximum likelihood estimation (MLE) was employed to follow previous research^20^. This factor analysis was conducted using oblique rotation (OBLIMIN), with the 176 question items entered as measurement variables. This data analysis used unscaled data to compute and analyze the heterogeneous correlation matrix because the data set included scales evaluated on a binary basis. All these procedures follow the method of Gillan et al. (2016), in which the authors extracted transdiagnostic dimensions using factor analysis. We separated large online samples into two balanced samples (Ds1: N=9753, Ds2: N=9752) and applied exploratory factor analysis in one dataset. We then did the same process in another one to confirm the validity of the result. The Cattel-Nelson-Gorsuch (CNG) test was performed in each dataset, and the optimal number of factors was extracted. Importantly, we used each factor score derived from factor analysis based on all samples for the following analysis.

#### Generalized mixed-effect linear model

Generalized linear mixed models were used to examine the association between each symptom and each diagnostic transdiagnostic dimension and BIS/BAS. Each symptom and transdiagnostic dimension was standardized and included as a dependent variable. Gender, age, occupation, and house income were controlled by being included in fixed effects, while each subject factor was included as a random effect. The equation is expressed as follows.

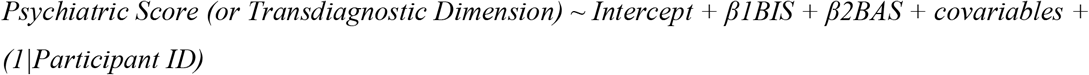

Factor analysis was conducted using R (4.2.3) and generalized linear regression using MatlabR2020b.

## Results

### Sample characteristics

The average age of the survey population was 41.5 years [standard deviation (SD) = 10.7], and 49.7% were male. Other details are described in Table 1.

**Table 1.** Characteristics of the survey population.

|  |  |  |  |  |  |  |  |  |  |  |
| --- | --- | --- | --- | --- | --- | --- | --- | --- | --- | --- |
| All | 19505<br>(100%) |  |  |  |  |  |  |  |  |  |
| Sex | Male | Female |  |  |  |  |  |  |  |  |
|  | 9687<br>(49.7%) | 9818<br>(50.4%) |  |  |  |  |  |  |  |  |
| Marital status | Not married | Married |  |  |  |  |  |  |  |  |
|  | 9427<br>(48.3%) | 10078<br>(51.7%) |  |  |  |  |  |  |  |  |
| The existence of child | No | Yes |  |  |  |  |  |  |  |  |
|  | 11015<br>(56.5%) | 8490<br>(43.5%) |  |  |  |  |  |  |  |  |
| Household income (yen) | Less than 2 million | 2 to Less than 4 million | 4 to Less than 6 million | 6 to less than 8 million | 8 to less than 10 million | 10 to less than 12 million | 12 to less than 15 million | 15 to less than 20 million | More than 20 million | Missing |
|  | 1525<br>(7.8%) | 3503<br>(18.0%) | 3729<br>(19.1%) | 2609<br>(13.4%) | 1599<br>(8.2%) | 777<br>(4.0%) | 400<br>(2.1%) | 217<br>(1.1%) | 119<br>(0.6%) | 5027<br>(25.8%) |
| Job | Employee or Executive | Self-employed | Homemaker | Other | No job |  |  |  |  |  |
|  | 10296<br>(52.8%) | 1224<br>(6.3%) | 2249<br>(11.5%) | 4529<br>(23.2%) | 1207<br>(6.2%) |  |  |  |  |  |

### Factor analysis to extract transdiagnostic dimensions

Our CNG test indicated three factors of latent structure (Figure 1a,b). The correlation between the two separated datasets was high in eigenvalues and each factor score (*r* > 0.9), indicating the structure’s robustness. We labeled each factor as ‘Compulsive hypersensitivity (CH),’ ‘Social withdrawal (SW),’ and ‘Addictive behavior (AB)’ based on the strongest individual item loadings.

**Figure 1.**
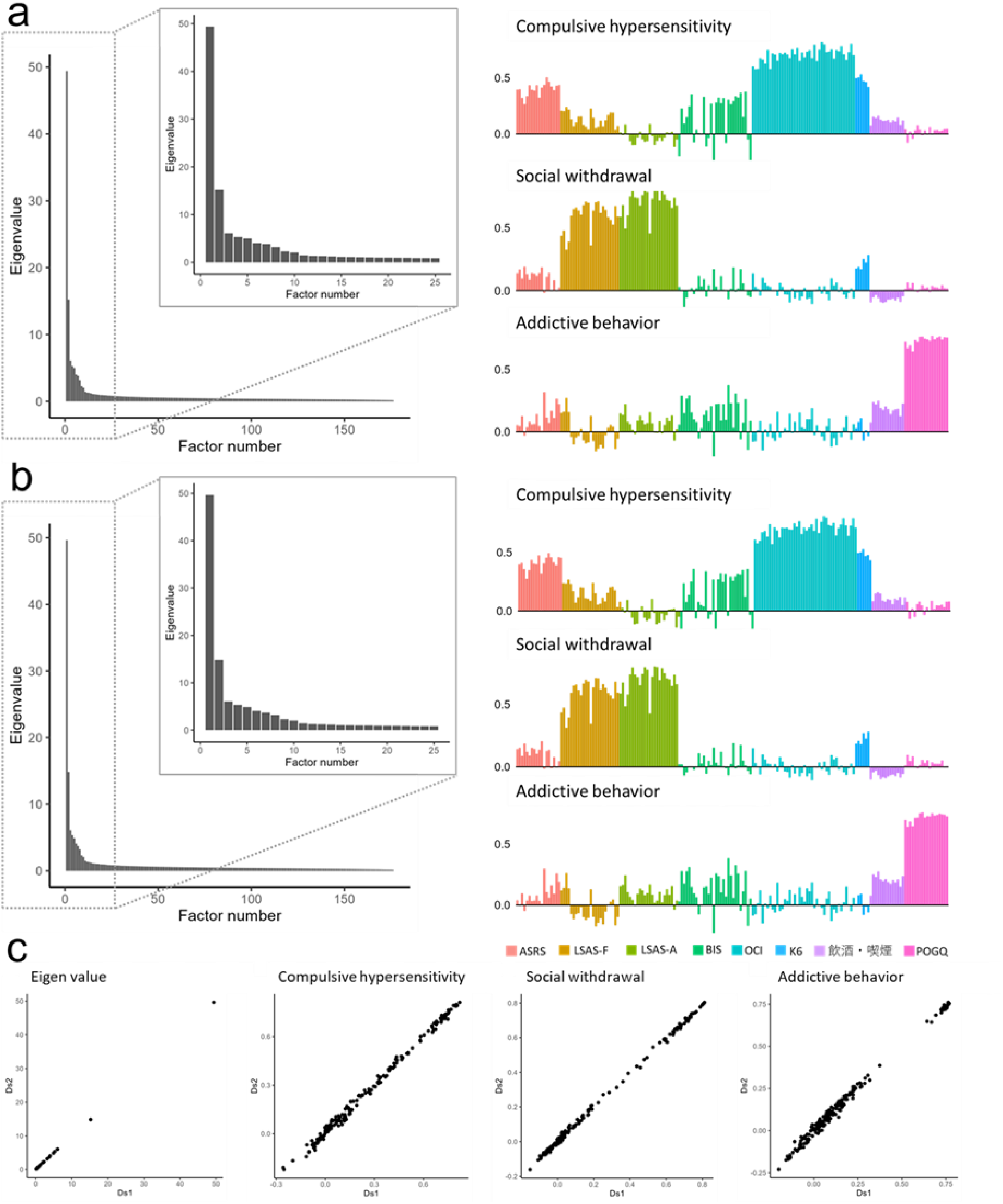
Extracting transdiagnostic factors. (**a**,**b**) Scree plot, correlation matrix, and item loadings from factor analysis in the separated datasets (Ds 1, Ds 2). The result suggested that the three-factor solution best explained the data from our samples. Factors were named ‘Compulsive hypersensitivity,’ ‘Social withdrawal,’ and ‘Addictive behavior.’ Each label meaning indicates the names of questionnaires as follows: ASRS attentional/hyperactivity disorder; LSAS-F/A social anxious symptoms of fear/avoidance; BIS Impulsivity; OCI obsessive-compulsive disorder; K6 Psychological distress; CAGE/TDS alcohol-related disorder and tobacco dependence; POGQ Gaming disorder. The reason why we use different colors for two factors of social anxiety separately and the same color for alcohol and tobacco use disorder is because the direction of loadings looks different/same in each factor. **(c) Scatter plots to compare the results from two datasets**. All correlations were more than r = 0.90, which indicates that the factor structure is robust.

For the CH factor, the highest average loadings came from OCD (Ds1: M=0.71, SD=0.06; Ds2: M=0.71, SD=0.06), followed by ADHD (Ds1: M=0.39, SD=0.06; Ds2: M=0.40, SD=0.06) and psychological distress (Ds1: M=0.48, SD=0.04; Ds2: M=0.49, SD=0.03). The highest average loadings of the second factor SW came from social anxiety (Ds1: M=0.65, SD=0.12; Ds2: M=0.64, SD=0.12), followed by psychological distress (Ds1: M=0.22, SD=0.05; Ds2: M=0.23, SD=0.04). For the third factor AB, the highest average loadings came from the gaming disorder (Ds1: M=0.73, SD=0.03; Ds2: M=0.72, SD=0.03), followed by alcohol-related disorder (Ds1: M=0.20, SD=0.08; Ds2: M=0.21, SD=0.09) and tobacco dependence (Ds1: M=0.19, SD=0.01; Ds2: M=0.20, SD=0.01). Notably, each transdiagnostic dimension included loading values from various items across questionnaires.

### Generalized mixed effect model revealed the relationship between psychopathological scores and reward/punishment sensitivity

Our regression analyses for each disorder indicated significant associations with reward/punishment sensitivity (Fig 2). With reward sensitivity, there were significant relationships in social anxiety (*β* = −0.061, Standard Error (SE) = 0.008, *p*_*FDR*_ < 0.001); gaming disorder (*β* = 0.076, SE = 0.008, *p*_*FDR*_ < 0.001); ADHD (*β* = 0.120, SE = 0.008, *p*_*FDR*_ < 0.001); OCD (*β* = 0.052, SE = 0.009, *p*_*FDR*_ < 0.001); PD (*β* = −0.030, SE = 0.008, *p*_*FDR*_ < 0.001); Tobacco dependence (*β* = 0.038, SE = 0.009, p_*FDR*_ < 0.001). With punishment sensitivity, there were significant relationships in social anxiety (*β* = 0.313, SE = 0.008, *p*_*FDR*_ < 0.001); gaming disorder (*β* = −0.076, SE = 0.009, *p*_*FDR*_ < 0.001); ADHD (*β* = 0.196, SE = 0.009, *p*_*FDR*_ < 0.001); OCD (*β* = 0.095, SE = 0.009, *p*_*FDR*_ < 0.001); PD (*β* = 0.318, SE = 0.008, *p*_*FDR*_ < 0.001); Alcohol-related disorder (*β* = −0.053, SE = 0.009, *p*_*FDR*_ < 0.001); Tobacco dependence (*β* = −0.037, SE = 0.009, *p*_*FDR*_ < 0.001). Impulsivity was not related to both sensitivities significantly.

**Figure 2.**
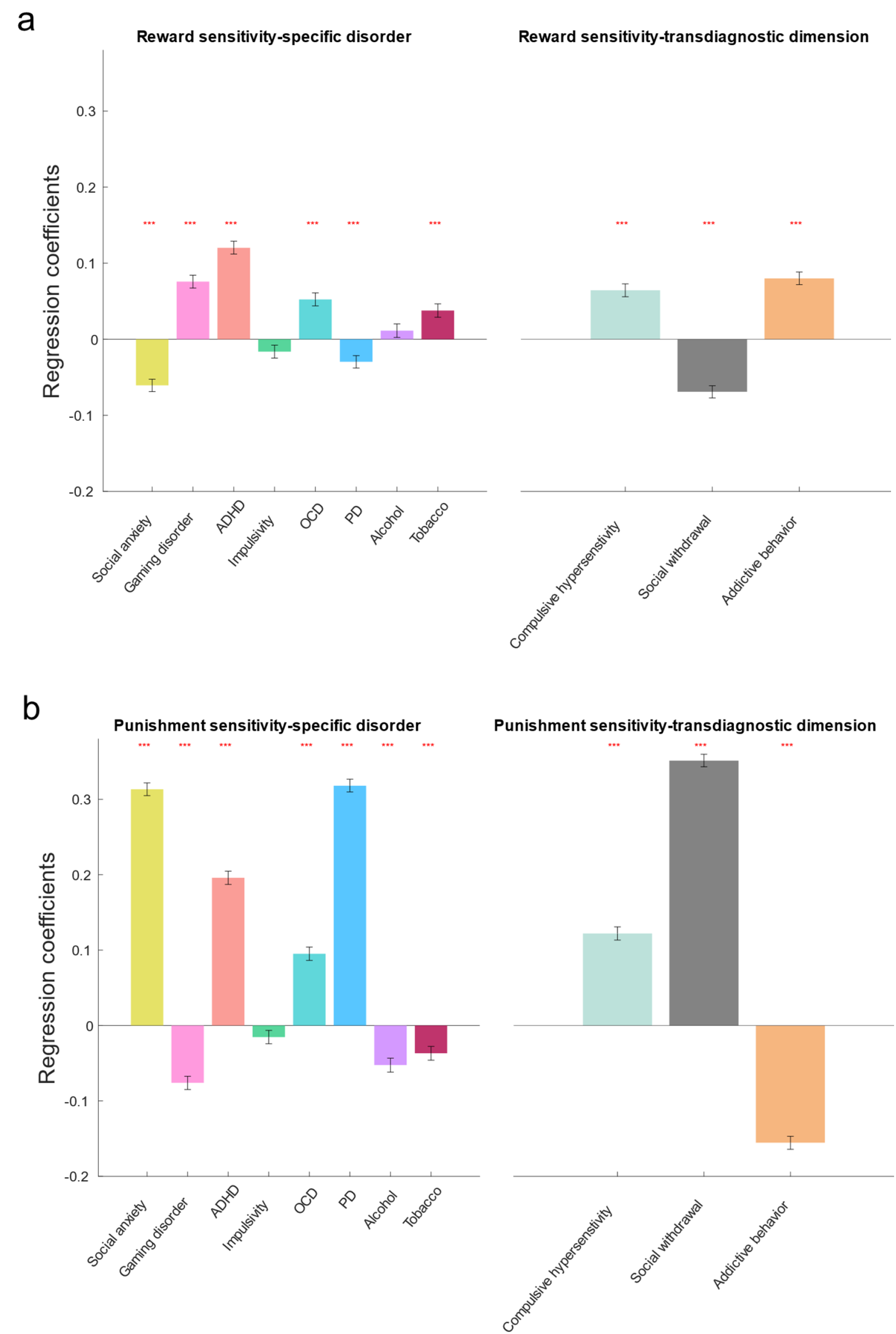
Associations between reward/punishment sensitivity and psychopathology. **(a)** The regression results for the relationship between reward sensitivity and psychopathology. **(b)** The regression results for the relationship between punishment sensitivity and psychopathology. ADHD attentional/hyperactivity disorder; OCD obsessive-compulsive disorder based; PD psychological distress. *** indicates p_FDR_ < 0.001.

Our regression analyses for transdiagnostic dimensions also revealed distinctive significant relationships between the factors. SW factor was associated with reward sensitivity negatively (β = −0.069, SE = 0.008, *p*_*FDR*_ < 0.001) and with punishment sensitivity positively (β = 0.351, SE = 0.008, *p*_*FDR*_ < 0.001). Contrarily, the AB factor was associated with reward sensitivity positively (β = 0.080, SE = 0.008, *p*_*FDR*_ < 0.001) and with punishment sensitivity negatively (β = −0.156, SE = 0.009, *p*_*FDR*_ < 0.001). Notably, CH was associated with both sensitivities positively, differently from the other two dimensions (β = 0.064, SE = 0.008, *p*_*FDR*_ < 0.001;β = 0.122, SE = 0.009, *p*_*FDR*_ < 0.001). See Table S1&S2 for the details of each statistical value.

## Discussion

Here, we used a dimensional approach to investigate the psychological mechanisms for transdiagnostic behavioral dysregulation and reward/punishment sensitivity in a large general population sample. Evidence from our analyses showed that the compulsive hypersensitivity factor is a symptom dimension associated with both reward and punishment, distinct from its relationships with the other two transdiagnostic dimensions. Interestingly, the compulsive hypersensitivity-related dimension includes a wide variety of items related to obsessiveness, impulsivity, inattention, and potential fear in social contexts.

With regard to our factor analysis, three robust transdiagnostic dimensions were extracted: ‘Compulsive hypersensitivity (CH),’ ‘Social withdrawal (SW),’ and ‘Addictive behavior (AB).’ Although there are differences from those obtained in past dimensional approaches^20,37^, given that the present study sought to capture psychopathological aspects characteristic focusing on behavioral dysregulation, it is natural that differences would arise with data-driven studies that also include more items related to general symptoms, e.g., major depression disorder. On the other hand, there are several commonalities, and the results of the study on compulsive hypersensitivity and social withdrawal are partially consistent with those of previous studies (e.g., compulsive and intrusive thoughts, social withdrawal^20,37^). These results suggest that transdiagnostic dimensions may be robust across regions, races, and cultures. Moreover, considering most existing research has not validated the factors in large hold-out samples, the fact that this study used approximately 10,000 samples as a hold-out group to confirm the robustness of the structure presents the validity of the transdiagnostic factors obtained here.

Our regression analyses found significant associations between categorical and transdiagnostic dimensions and reward/punishment sensitivity, respectively. Significant negative and positive associations existed between SW and reward/punishment sensitivity, respectively. Similarly, there were positive and negative associations between AB and these sensitivities. These findings align with previous studies^38,39^ exploring the relationships from a transdiagnostic perspective. However, even though the association of the scores of alcohol-related disorder and impulsivity with reward sensitivity was not significant when viewed by categorical disorder, the fact that the relationship with AB was significant suggests that the association could have been found more robustly based on an index based on transdiagnostic symptom dimensions. The specificity of CH, which is constructed based on OCD and ADHD transdiagnostically, is associated with greater sensitivity to both reward and punishment in this dimension, unlike the other two dimensions. The results support a critique of the notion that compulsivity and impulsivity are at opposite ends of the approach/avoidance spectrum. They suggest that compulsive-impulsive dyscontrol may coexist at a higher level, as many clinicians and researchers have proposed regarding reward/punishment processing^40,41^.

On the other hand, reward/punishment treatment was assessed based on subjective reports; therefore, its ecological validity is questionable compared to behavioral indexes and parameters derived from computational modeling. Indeed, such self-reported indexes may not be directly related to behaviorally or computationally defined reward/punishment sensitivity^42^. However, this does not simply mean that such a subjective index does not have validity. It also indicates that it is a psychological characteristic captured by the individual’s subjective awareness and that the layer may differ from the behavioral and computational one. In fact, studies that have investigated the relationship between psychopathology, behavioral measures, and subjective self-esteem have also suggested that self-esteem as a higher-level concept that sits above behavioral measures best explains psychopathology^37^. Considering the assumption of such complex relationships, future research should clarify the relationship between subjective sensitivity and psychopathology and reward/punishment sensitivity, which should be formulated through a combination of behavioral experiments and computational modeling^43^.

Moreover, it should be noted that even though traits such as reward/punishment sensitivity examine psychopathology, the characteristics are not pathology per se. In addition, the association between psychopathology and reward/punishment sensitivity might be mediated by other neuropsychological factors such as meta-cognition^44^. When considering more general applications, such as psychotherapy, a transdiagnostic dimensional model should be used to identify detailed mediating processes, leading to an approach that increases the precision of intervention targeting. Through such detailed future research, the transdiagnostic dimensional model allows us to take a more granular approach rather than merely focusing on categorical psychiatric disorders.

Given the complexity of the association between behavioral dysregulation and various psychological constructs, this study has several limitations. First, as this study is cross-sectional, it does not shed light on time-dependent and causal associations. Considering the recent suggestion that psychiatry needs time and context^45,46^, future research should apply a longitudinal approach and take care of spatiotemporal aspects. Second, the psychiatric scores assessed here do not include essential dimensions for behavioral dysregulation, such as eating disorders. However, it has been reported that general factors of psychopathology are identifiable, even if the measures assessing symptoms do not cover all elements^47^, which means this issue is negligible to some extent here. Third, our analysis focuses on the Japanese population. A deeper understanding of the transdiagnostic psychological mechanisms of behavioral dysregulation would benefit from international comparisons including various races, cultures, religions, and other statuses, which were not assessed in the present study.

Fourth, these surveys were conducted using online recruitment methods, which may introduce sampling bias. Generalized linear model analyses corrected for this possibility by adjusting for confounding factors such as age, gender, and other socio-demographic attributes, reducing the influence of bias.

In summary, the symptoms from eight psychiatric disorders related to compulsive-impulsive behavioral dysregulation were aggregated into three factors with different relationships with reward/punishment sensitivity. Our findings underline the association between compulsive hypersensitivity and reward and punishment sensitivities. This finding highlights the importance of a transdiagnostic multidimensional approach for examining these relationships with psychological aspects. Furthermore, our results contribute to a step forward toward establishing a transdiagnostic framework, such as the RDoC, and suggest that dimensional symptoms of psychiatric disorders may correspond more clearly to psychological constructs than to existing overlapping and heterogeneous definitions of psychiatric disorders.

## Data Availability

The manuscript contains all summary statistics supporting this study's findings. The factor loadings of each transdiagnostic dimension will be accessible in the Open Science Framework (OSF). Owing to company cohort data sharing restrictions, individual-level data cannot be publicly posted. However, data are available from the authors upon request and with the permission of KDDI Corporation.

## Declaration of AI use

We have used AI-assisted technologies to improve the manuscript’s readability and language.

## Acknowledgment

This research was supported by the KDDI collaborative research contract and JSPS KAKENHI grant number 24K22822 (TO).

## Author contributions

TO designed the study and prepared the original draft; TO, AS, and NK acquired and analyzed the data; AS and NK reviewed and edited the manuscript. All authors gave final approval for submission and agreed to take responsibility for the manuscript.

