## Supplementary materials for "A Transdiagnostic Dimensional Approach to Behavioral Dysregulation: Examining Reward and Punishment Sensitivity Across Psychopathology"

### Supplementary Results

| | Reward sensitivity | | | Punishment sensitivity | | |
|---|---|---|---|---|---|---|
| | Coef | SE | *pFDR* | Coef | SE | *pFDR* |
| **Social anxiety** | -0.061 | 0.008 | <0.001 | 0.313 | 0.008 | <0.001 |
| **Gaming disorder** | 0.076 | 0.008 | <0.001 | -0.076 | 0.009 | <0.001 |
| **ADHD** | 0.120 | 0.008 | <0.001 | 0.196 | 0.009 | <0.001 |
| **Impulsivity** | -0.016 | 0.009 | 0.056 | -0.015 | 0.009 | 0.082 |
| **OCD** | 0.052 | 0.009 | <0.001 | 0.095 | 0.009 | <0.001 |
| **PD** | -0.030 | 0.008 | <0.001 | 0.318 | 0.008 | <0.001 |
| **Alcohol** | 0.011 | 0.009 | 0.203 | -0.053 | 0.009 | <0.001 |
| **Tobacco** | 0.038 | 0.009 | <0.001 | -0.037 | 0.009 | <0.001 |

**Table S1 The regression results for the relationship between reward sensitivity and psychopathology**

*ADHD attentional/hyperactivity disorder; OCD obsessive-compulsive disorder based; PD psychological distress.*

| | Reward sensitivity | | | Punishment sensitivity | | |
|---|---|---|---|---|---|---|
| | Coef | SE | *pFDR* | Coef | SE | *pFDR* |
| **Compulsive hypersensitivity** | 0.064 | 0.008 | <0.001 | 0.122 | 0.009 | <0.001 |
| **Social withdrawal** | -0.069 | 0.008 | <0.001 | 0.351 | 0.008 | <0.001 |
| **Addictive behavior** | 0.080 | 0.008 | <0.001 | -0.156 | 0.009 | <0.001 |

**Table S2 The regression results for the relationship between punishment sensitivity and psychopathology.**
